# The Effect of Convalescent Plasma Therapy on COVID-19 Patient Mortality: Systematic Review and Meta-analysis

**DOI:** 10.1101/2020.07.29.20162917

**Authors:** Stephen A. Klassen, Jonathon W. Senefeld, Patrick W. Johnson, Rickey E. Carter, Chad C. Wiggins, Shmuel Shoham, Brenda J. Grossman, Jeffrey P. Henderson, James Musser, Eric Salazar, William R. Hartman, Nicole M. Bouvier, Sean T. H. Liu, Liise-anne Pirofski, Sarah E. Baker, Noud van Helmond, R. Scott Wright, DeLisa Fairweather, Katelyn A. Bruno, Zhen Wang, Nigel S. Paneth, Arturo Casadevall, Michael J. Joyner

## Abstract

To determine the effect of COVID-19 convalescent plasma on mortality, we aggregated patient outcome data from 10 randomized clinical trials (RCT), 20 matched-control studies, two dose-response studies, and 96 case-reports or case-series. Studies published between January 1, 2020 – January 16, 2021 were identified through a systematic search of online PubMed and MEDLINE databases. Random-effects analyses of RCT and matched-control data demonstrated that COVID-19 patients transfused with convalescent plasma exhibited a lower mortality rate compared to patients receiving standard treatments. Additional analyses showed that early transfusion (within 3 days of hospital admission) of higher-titer plasma is associated with lower patient mortality. These data provide evidence favoring the efficacy of human convalescent plasma as a therapeutic agent in hospitalized COVID-19 patients.

## Introduction

Convalescent plasma is a century-old passive antibody therapy that has been used to treat outbreaks of novel infectious diseases, including those affecting the respiratory system.^1,2^ At the onset of the pandemic, human convalescent plasma was used world-wide as it represented the only antibody-based therapy to treat coronavirus disease 2019 (COVID-19), caused by the severe acute respiratory syndrome coronavirus 2 (SARS-CoV-2).^2–5^ Despite the emerging availability of monoclonal antibody therapies and vaccines for use in non-hospitalized patients via federal emergency authorization routes, convalescent plasma usage has persisted (∼100,000 units/month in the United States in early 2021) during subsequent waves of the COVID-19 pandemic due to surging hospitalizations and mortality rates.^6–9^ However, evidence for therapeutic COVID-19 convalescent plasma efficacy still requires definitive support from large randomized clinical trials (RCT). As a result, there remains a lack of consensus on convalescent plasma use in hospitalized COVID-19 patients.^10,11^ Smaller RCTs, matched-control studies, and case-series studies investigating convalescent plasma therapy for COVID-19 have emerged and provided a positive efficacy signal.^12–18^ Most of these studies, however, lacked appropriate statistical power or were terminated early. Also, many studies have transfused patients only after clinical progression to severe COVID-19 respiratory distress, which opposes historical data highlighting the efficacy of early convalescent plasma transfusion and overlooks viral neutralization as the fundamental mechanism for convalescent plasma therapy.^1,2^

There is an urgent need to determine the efficacy of potential treatments amidst the ongoing COVID-19 pandemic. Although a ‘living’ systematic review has summarized a broad-ranging clinical experience with convalescent plasma,^10,11^ this approach may be limited because it employed stringent inclusion criteria for aggregating patient outcomes, which prevented a preliminary assessment of convalescent plasma efficacy. Given the insufficient patient outcome data available from RCTs, we used a pragmatic approach for study selection to aggregate COVID-19 clinical outcomes, focusing solely on mortality data from RCTs, matched-control studies, dose-response investigations, and case-series or case reports in real time. Our primary objective was to derive an aggregate estimate of the mortality rates from transfused and non-transfused cohorts of contemporaneous COVID-19 studies. As an exploratory objective, we assessed whether the time from hospital admission to convalescent plasma transfusion was associated with patient mortality.

## Methods

### Eligibility

We included RCTs, matched-control trials, dose-response studies, and case-series or case-reports published on pre-print servers or peer-reviewed journals that investigated the impact of human convalescent plasma therapy on COVID-19 patient mortality.

### Literature Search and Data Extraction

We performed a systematic search of the online Pubmed and MEDLINE databases from January 1, 2020 through January 16, 2021. Keywords used in the search included: ((convalescent plasma) OR (convalescent serum)) AND COVID-19 (and medical subject headings; MeSH terms) using the following limits: Humans. No language restrictions were imposed. The references of all eligible studies were reviewed to identify other potentially eligible studies. In order to be considered eligible for inclusion, studies must have: 1) included patients with confirmed diagnosis of COVID-19, 2) used convalescent plasma treatment, and 3) reported mortality. Randomized clinical trials, matched-control studies, dose-response studies, case-series and case-reports were included. Two reviewers (S.A.K and J.W.S) independently screened the titles and abstracts of all studies identified by the search to determine eligibility. Studies that were deemed potentially eligible had their full text reviewed (S.A.K and J.W.S) in order to determine if they met the criteria for inclusion in the review. Disagreement was resolved by consensus. Two reviewers (S.A.K and J.W.S) extracted study and patient characteristics as well as clinical information (variables delineated in **Supplemental Tables S4, S5, S6**).

Two reviewers (S.A.K and J.W.S) independently assessed the risk of bias for mortality data of each included study using the Cochrane Risk of Bias criteria (for RCTs) and the Newcastle-Ottawa Scale (for matched-control studies) (**Supplement tables S1 and S2**).^19–21^ Dose-response studies were evaluated with the Newcastle-Ottawa Scale. The criteria developed by the Mayo Clinic Evidence-Based Practice Research Program informed our assessment of bias in the mortality data reported by case-series and case-reports.^22^

### Data Synthesis

For RCTs and matched-control studies, we recorded the number of survivors and non-survivors in transfused and non-transfused cohorts to calculate odds ratios with 95% confidence intervals. For dose-response studies, we recorded the number of survivors and non-survivors among patients who were transfused with higher-titer and lower-titer convalescent plasma unit sub-groups to calculate odds ratios with 95% confidence intervals. Aggregate mortality rates were calculated for transfused and, if applicable, non-transfused patients at the longest reported vital status for each study.

Using the DerSimonian–Laird random-effects method^23^ we computed aggregate odds ratios with 95% confidence intervals separately for RCTs and matched-control studies. We also computed aggregate odds ratios with 95% confidence intervals for RCTs and matched-control studies, combined. Simple random-effects meta-regression analyses evaluated the moderator variables (i.e., cohort age, proportion of cohort receiving mechanical ventilation, and duration of study follow up) on mortality for all clinical studies. The I^2^ statistic was used to quantify heterogeneity. Based on historical data,^1^ we performed an exploratory subgroup analysis to assess the impact of early transfusion (within 3 days of hospital admission) compared to late transfusion (>3 days after hospital admission) on COVID-19 patient mortality. All analyses were performed with Comprehensive Meta-analysis Software (Biostat, version 3.3.070). Tests were two-tailed and alpha was 0.05. Figures were made with R software (R Core Team). The number needed to treat was calculated using aggregate data from controlled studies.^24^ Dose-response studies, case-series and case-reports were not included in the meta-analysis but were described in a narrative.

### Certainty of Evidence Assessment

We used the GRADE approach to assess the certainty of evidence regarding the impact of convalescent plasma on COVID-19 patient mortality.^25^ The risk of bias assessments for RCT and matched-control data informed our certainty of evidence assessment.

## Results

### Search Results

The literature search yielded 780 studies, of which 128 studies met the eligibility criteria and were included in the systematic review (**Supplemental Figure S1**). The present analyses included a total of 10 RCTs,^13,18,26–33^ 20 matched-control studies,^16,34–53^ two dose-response studies,^54,55^ and 96 case-series or case-reports.^3,15,40,46,56–147^ Overall, these studies reported outcomes from 35,055 COVID-19 patients in 31 countries (**Table 1, Table 2, and Supplemental Table S3**). The age of patients enrolled in these studies ranged from 4 to 100 years, with a greater proportion of men than women in most studies (proportion of women: 0% to 100%) (**Supplemental Tables S4, S5, S6**). All studies included patients with diagnosed COVID-19, with most studies including hospitalized patients with severe or life-threatening COVID-19. At the time of plasma transfusion, the proportion of patients on mechanical ventilation varied by study from 0% to 100%. The duration of follow up ranged from 2 to 118 days (**Supplemental Tables S4, S5, S6**). In most studies, patients were eligible to receive concomitant and experimental therapies such as antivirals, steroids, and chloroquine or hydroxychloroquine.

**Table 1.** Mortality Rates among COVID-19 Patients.

| Table 1 \| Mortality Rates among COVID-19 Patients |  |  |  |  |  |  |  |  |  |  |
| --- | --- | --- | --- | --- | --- | --- | --- | --- | --- | --- |
| Study | Location | Convalescent Plasma |  |  | Control |  |  | Statistics |  |  |
|  |  | Survivor | Non-Survivor | Mortality | Survivor | Non-Survivor | Mortality | OR | P | 95% CI |
| Randomized Clinical Trials |  |  |  |  |  |  |  |  |  |  |
| Avendano-Sola et al. | ESP | 38 | 0 | 0% | 39 | 4 | 9% | 0.11 | 0.15 | 0.01, 2.19 |
| Rasheed et al. | IRQ | 20 | 1 | 5% | 20 | 8 | 29% | 0.13 | 0.06 | 0.01, 1.09 |
| Gharbharan et al. | NLD | 37 | 6 | 14% | 32 | 11 | 26% | 0.47 | 0.18 | 0.16, 1.42 |
| AlQahtani et al. | BHR | 19 | 1 | 5% | 18 | 2 | 10% | 0.47 | 0.56 | 0.04, 5.69 |
| Libster et al. | ARG | 78 | 2 | 3% | 76 | 4 | 5% | 0.49 | 0.41 | 0.09, 2.74 |
| Li et al. | CHN | 43 | 8 | 16% | 38 | 12 | 24% | 0.59 | 0.30 | 0.22, 1.59 |
| Ray et al. | IND | 30 | 10 | 25% | 26 | 14 | 35% | 0.62 | 0.33 | 0.24, 1.63 |
| Simonovich et al. | ARG | 197 | 25 | 11% | 93 | 12 | 11% | 0.98 | 0.96 | 0.47, 2.04 |
| Agarwal et al. | IND | 201 | 34 | 14% | 198 | 31 | 14% | 1.08 | 0.77 | 0.64, 1.83 |
| Bajpai et al. | IND | 11 | 3 | 21% | 14 | 1 | 7% | 3.82 | 0.27 | 0.35, 41.96 |
| Random Effects Model |  | 674 | 90 | 12% | 554 | 99 | 15% | 0.76 | 0.14 | 0.54, 1.09 |
| Random Effects Model excluding Agarwal et al. |  | 473 | 56 | 11% | 356 | 68 | 16% | 0.65 | 0.04 | 0.43, 0.98 |
| Matched-Control Studies |  |  |  |  |  |  |  |  |  |  |
| Duan et al. | CHN | 10 | 0 | 0% | 7 | 3 | 30% | 0.10 | 0.15 | 0.01, 2.28 |
| Perotti et al. | ITA | 43 | 3 | 7% | 16 | 7 | 30% | 0.16 | 0.01 | 0.04, 0.69 |
| Omrani et al. | QAT | 39 | 1 | 3% | 35 | 5 | 13% | 0.18 | 0.13 | 0.02, 1.61 |
| Hegerova et al. | Washington, USA | 18 | 2 | 10% | 14 | 6 | 30% | 0.26 | 0.13 | 0.05, 1.49 |
| Salazar E. et al. | Texas, USA | 146 | 6 | 4% | 235 | 34 | 13% | 0.28 | 0.01 | 0.12, 0.69 |
| Alsharidah et al. | KWT | 111 | 24 | 18% | 143 | 90 | 39% | 0.34 | <0.001 | 0.21, 0.57 |
| Zeng Q. et al. | CHN | 1 | 5 | 83% | 1 | 14 | 93% | 0.36 | 0.50 | 0.02, 6.85 |
| Donato et al. | New York, USA | 36 | 11 | 23% | 775 | 565 | 42% | 0.42 | 0.01 | 0.21, 0.83 |
| Salazar M. et al. | ARG | 647 | 221 | 25% | 1288 | 1010 | 44% | 0.44 | <0.001 | 0.37, 0.52 |
| Liu et al. | New York, USA | 34 | 5 | 13% | 118 | 38 | 24% | 0.46 | 0.13 | 0.17, 1.25 |
| Xia et al. | CHN | 135 | 3 | 2% | 1371 | 59 | 4% | 0.52 | 0.27 | 0.16, 1.67 |
| Abolghasemi et al. | IRN | 98 | 17 | 15% | 56 | 18 | 24% | 0.54 | 0.10 | 0.26, 1.13 |
| AlShehry et al. | SAU | 30 | 10 | 25% | 78 | 46 | 37% | 0.57 | 0.16 | 0.25, 1.26 |
| Budhiraja et al. | IND | 248 | 85 | 26% | 241 | 120 | 33% | 0.69 | 0.03 | 0.50, 0.96 |
| ah Yoon et al. | New York, USA | 50 | 23 | 32% | 45 | 28 | 38% | 0.74 | 0.39 | 0.37, 1.46 |
| Rogers et al. | Rhode Island, USA | 56 | 8 | 13% | 149 | 28 | 16% | 0.76 | 0.52 | 0.33, 1.77 |
| Altuntas et al. | TUR | 669 | 219 | 25% | 642 | 246 | 28% | 0.85 | 0.15 | 0.69, 1.06 |
| Klapholz et al. | New Jersey, USA | 37 | 10 | 21% | 38 | 9 | 19% | 1.14 | 0.80 | 0.42, 3.13 |
| Klein et al. | Maryland, USA | 25 | 9 | 26% | 26 | 8 | 24% | 1.17 | 0.78 | 0.39, 3.51 |
| Moniuszko-Malinowska et al. | POL | 49 | 6 | 11% | 672 | 43 | 6% | 1.91 | 0.16 | 0.78, 4.72 |
| Random Effects Model |  | 2482 | 668 | 21% | 5950 | 2377 | 29% | 0.57 | <0.001 | 0.45, 0.72 |
| Overall Random Effects Model <sup>a</sup> |  | 2955 | 724 | 20% | 6306 | 2445 | 28% | 0.58 | <0.001 | 0.47, 0.71 |

CI, confidence interval; OR, odds ratio
<sup>a</sup> random-effects model excludes trial by Agarwal et al.
CI, confidence interval; OR, odds ratio
<sup>a</sup> random-effects model excludes trial by Agarwal et al.

**Table 2.**
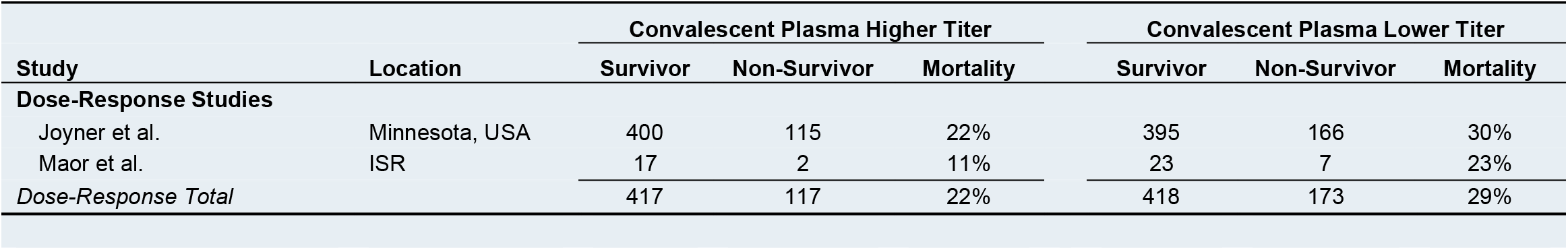
Mortality Rates among COVID-19 Patients.

### Meta-analysis

#### Randomized Clinical Trials

When data from the 10 RCTs were aggregated, there was no association between convalescent plasma therapy and mortality (odds ratio [OR]: 0.76, 95% confidence interval [95% CI]: 0.54, 1.09, *P* = 0.14, *I*^*2*^ *=* 7%) (**Table 1 and Figure 1**). Although the heterogeneity was low, one RCT (Agarwal et al.^32^) demonstrated a directionally different effect, had a large statistical weight (34.2), and represented the primary source of heterogeneity (Δ*I*^*2*^ = 7%). Additionally, in the context of COVID-19, neutralizing antibodies are hypothesized to represent the primary active agent in convalescent plasma and the marker of plasma potency.^148,149^ In this regard, as mentioned below, two studies report a dose-response relationship between convalescent plasma antibody level and mortality, suggesting the need for a sufficient amount of antibody for therapeutic success.^148,149^ The Agarwal et al.^32^ trial included a large proportion of patients (∼70%) in the convalescent plasma arm whom received plasma with low levels of SARS-CoV-2 antibodies less than 1:80, with ∼30% receiving plasma with no detectable antibodies.^32^ Thus, there were strong analytical and biological rationales to exclude this study from statistical models.

**Figure 1.**
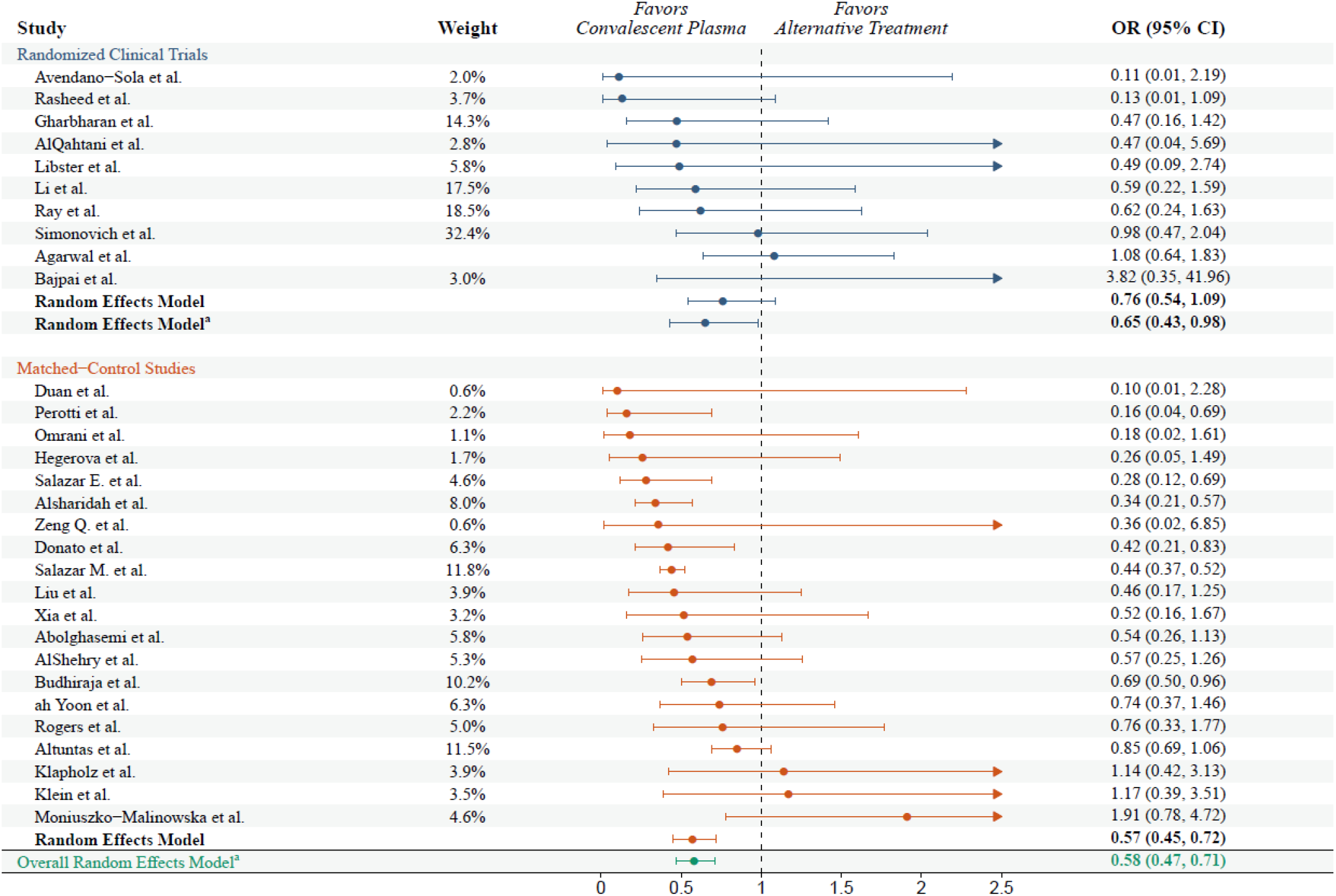
The effect of human convalescent plasma therapy on COVID-19 patient mortality. Forest plot illustrating odds ratios and 95% confidence intervals computed for each study and aggregated for each study type (DerSimonian–Laird random-effects model). Data are separated by study type with randomized clinical trials presented in blue and matched-control studies presented in orange. Odds ratios for each study type are presented in darker hues and the overall odds ratio pooled across all controlled studies is presented in green. Relative study weights are provided. The *I*^*2*^ values were 0 (randomized clinical trial model), 61 (matched-control study model), and 53 (overall model combining randomized clinical trial and matched-control studies). CI, confidence interval; OR, odds ratio. ^**a**^ Random-effects model excludes trial by Agarwal and colleagues

When analyses were performed on data from nine RCTs excluding the Agarwal et al.^32^ study, patients transfused with convalescent plasma exhibited a lower mortality rate compared to non-transfused COVID-19 patients (11% vs. 16% mortality; OR: 0.65, 95% CI: 0.43, 0.98, *P* = 0.04, *I*^*2*^ = 0%) (**Table 1 and Figure 1**). The aggregate OR (0.65) indicates that convalescent plasma was associated with a 35% reduction in the odds of mortality among COVID-19 patients.

#### Matched-control studies

When we aggregated mortality data from the 20 matched-control studies, patients transfused with convalescent plasma exhibited a lower mortality rate compared to non-transfused patients (21% vs. 29% mortality; OR: 0.57, 95% CI: 0.45, 0.72, *P* < 0.001, *I*^*2*^ = 61%) (**Table 1 and Figure 1**).

#### Randomized Clinical Trials and Matched-control studies

Aggregation of mortality data from all controlled studies including RCTs and matched-control studies indicated that patients transfused with convalescent plasma exhibited a 42% reduction in mortality rate compared to patients receiving standard treatments (20% vs. 28% mortality; OR: 0.58, 95% CI: 0.47, 0.71, *P* < 0.001, *I*^*2*^ = 53%) (**Table 1 and Figure 1**). Simple random-effects meta-regression analyses indicated that cohort age (*P* = 0.23), proportion of cohort receiving mechanical ventilation (*P* = 0.51), and duration of study follow up (*P* = 0.29) did not affect the aggregate OR computed for all controlled studies.

#### Subgroup analysis: effect of days between hospital admission and plasma transfusion

Sixteen studies (*n* = 6 RCTs, *n* = 10 matched-control studies) reported the number of days between hospital admission and convalescent plasma transfusion (**Supplemental Table S4**). Exploratory analysis revealed that the mortality reduction associated with convalescent plasma transfusion was greater in studies that transfused patients within 3 days of hospital admission (OR: 0.44 95% CI: 0.32, 0.61) compared to studies that transfused patients >3 days after hospital admission (OR: 0.79 95% CI: 0.62, 0.98; Random effects test of heterogeneity between subgroups: *P* = 0.005). However, this analysis was strongly influenced by the study by Altuntas and colleagues^42^ which transfused patients >3 days after admission (relative weight: 73%). Upon removing the study by Altuntas and colleagues,^42^ the number of days from hospital admission to transfusion no longer affected the mortality reduction associated with convalescent plasma transfusion (transfusion within 3 days of hospitalization: 0.44 [0.32, 0.60]; transfusion >3 days after hospitalization: 0.61 [0.36, 0.68]; Random effects test of heterogeneity between subgroups: *P* = 0.23).

### Additional Evidence

#### Dose-response studies

Two studies investigated the association between convalescent plasma antibody levels and the risk of mortality from COVID-19.^3,55^ Although different criteria were used to categorize convalescent plasma units as higher and lower antibody level, both studies found a dose-response association between antibody level and COVID-19 morality, such that patient mortality was lower in the subgroups transfused with higher-titer plasma. The aggregate mortality rate of COVID-19 patients transfused with higher-titer convalescent plasma was lesser than patients transfused with lower-titer plasma (22% vs. 29% mortality).

#### Case-series and case-reports

The aggregate mortality rate among COVID-19 patients transfused with convalescent plasma reported in uncontrolled studies was 13% (range: 0% to 100%), which is comparable to the mortality rates exhibited by transfused cohorts from clinical trials and matched-control studies (**Supplemental Table S3**). Case-series and case-report data included diverse patient cohorts with varying inherent risk for COVID-19 complications. Several studies explored immunosuppressed patients with suppressed antibody production due to hematological malignancies, cancer-directed therapy, or X-linked agammagloblulinemia (XLA) and provided an important ‘experiment of nature’ to evaluate convalescent plasma efficacy for COVID-19.^95,127,150,151^ For example, Jin et al.^95^ highlighted a series of three XLA patients with severe COVID-19 that failed to respond to other supportive treatments but demonstrated strong improvements in oxygen requirements and viral clearance within days of receiving convalescent plasma transfusions.

#### Risk of Bias

Overall, we deemed the risk of bias for mortality data to be low-to-moderate for RCTs and low-to-moderate for matched-control studies. We present the full judgement for each study in **Supplemental Table S1 and S2**. The risk of bias for uncontrolled studies is inherently high. Visual inspection of the funnel plot to assess publication bias shows that one study falls below the 95% CI and two studies fall above the 95% CI (**Supplemental Figure S2**). The funnel plot shows symmetry in the effect sizes among studies with low standard error and asymmetry among studies with greater standard error, suggesting that smaller studies with larger standard error may be more likely to report an effect of convalescent plasma. However, Egger’s regression test suggests that there is no significant asymmetry of the plot (intercept = −0.17, *P* = 0.67).

#### Certainty of Evidence

The certainty in the estimate of the effect of convalescent plasma on mortality is moderate-to-high.^152^ This judgment was based on the consistency of the results between RCT and matched-control studies and the corroborating evidence from dose-response studies and other uncontrolled case data. When aggregating data from all controlled studies, the meta-analyses provided precise estimates, did not demonstrate substantial heterogeneity, and demonstrated no strong evidence of publication bias. The inherent limitations of the included studies rendered the certainty of evidence judgement to be moderate-to-high.

#### Number needed to treat

Based on the aggregate OR (0.58, 95% CI: 0.47, 0.71) computed for all controlled studies and the aggregate mortality rate (28%) expressed by non-transfused cohorts among the controlled studies, in order to avoid one death the number needed to be transfused with convalescent plasma rather than only receive the standard of care is 11 (range: 8 – 16).

## Discussion

This analysis represents the most current aggregation of mortality data from contemporaneous COVID-19 convalescent plasma studies. The aggregate mortality rate of transfused COVID-19 patients was lower than that of non-transfused COVID-19 patients. Additional analyses demonstrated that early transfusion of high-titer plasma reduces mortality among patients with COVID-19. These results favor the efficacy of convalescent plasma as a COVID-19 therapeutic agent. The primary biological hypothesis for the efficacy of convalescent plasma is antibody-mediated SARS-CoV-2 viral neutralization and interference with viral replication, though other biological mechanisms may also contribute to the mitigation of symptoms.^2^ The mortality reduction associated with convalescent plasma aligns with similar analyses of historical data from convalescent plasma trials for viral diseases such as the 1918 flu epidemic,^1^ severe acute respiratory syndrome,^153^ and H1N1 influenza.^154^ Our findings are discordant with those of a previous ‘living’ systematic review,^10,11^ which concluded that there was insufficient evidence to determine the impact of convalescent plasma on all-cause mortality based on only two RCTs, including one prematurely terminated RCT (Li et al.^12^). This discordance reflects differences in the studies included in the analysis. Our approach was pragmatic and used less stringent study inclusion criteria, allowing for the inclusion of 30 controlled studies, of which a majority found a directionally similar effect of convalescent plasma and our analyses stratified by study design (e.g., RCT and matched-control studies) revealed similar aggregate ORs.

Mechanistic and clinical data support the reduction in mortality associated with convalescent plasma administration. Importantly, convalescent plasma contains SARS-CoV-2 neutralizing antibodies.^155,156^ Convalescent plasma administration increases SARS-CoV-2 clearance in COVID-19 patients^12,34^ including immunocompromised individuals,^95,110,121,150^ indicating an antiviral effect. Viral neutralization is then posited to reduce the inflammatory response and thus lessen the likelihood that an over-exuberant immune response progresses to lung damage, interference with gas exchange, and death. Additional evidence arising from animal studies shows that administration of human convalescent plasma is protective against SARS-CoV-2 infection.^157,158^ Antibody-mediated interference with viral replication may increase tissue repair and eventually manifest as reduced mortality. In addition, convalescent plasma transfusion is associated with reductions in inflammatory markers, such as chemokines, cytokines and C-reactive protein.^127,159^ Concomitant reductions in inflammation and improved gas exchange may underlie the reductions in oxygen requirements associated with convalescent plasma, even in critically ill patients. These findings provide mechanistic evidence for the reduction in mortality observed in patients receiving convalescent plasma.

There are several limitations to this analysis including aggregating mortality data across study populations that varied by: 1) the nation of data origin, 2) timing relative to world-wide progression of the pandemic, 3) clinical diagnostic and treatment algorithms, 4) plasma antibody titer and administration volume, 5) the latency between COVID-19 diagnosis and transfusion, and 6) the duration of follow up after transfusion. Also, we did not consult a librarian when constructing our search terms. However, high-quality evidence from large RCTs remains unavailable and the continuing global health emergency related to COVID-19 necessitated a practical real-time aggregation of existing mortality data. We note that the reports cited herein include positive results from different countries, suggesting that efficacy is robust across different health systems. Given the safety of convalescent plasma administration in COVID-19 patients,^3,4^ the results of this real-time systematic review and meta-analysis provide encouragement for its continued use as a therapy and may have broad implications for the treatment of COVID-19 and design of RCTs. Importantly, many of the patients enrolled in the studies included in the present analyses received convalescent plasma transfusions later in their disease course. In this context, prior to antibiotics and effective vaccinations, convalescent plasma therapy was widely understood to be most efficacious very early in the course of hospitalizations.^2,160^ As a result, our analysis may underestimate the mortality reduction achievable through early administration of high-titer convalescent plasma for COVID-19.

## Conclusion

This real-time systematic review and meta-analysis of contemporaneous studies highlights that the mortality rate of transfused COVID-19 patients was lower than that of non-transfused COVID-19 patients and suggests that early transfusion of high-titer plasma represents the optimal use scenario to reduce the risk of mortality among patients with COVID-19. These results favor the efficacy of convalescent plasma as a COVID-19 therapeutic agent.

## Data Availability

All procedures accessed public information as sourced in the main text.

## Acknowledgements

The authors express their gratitude to convalescent plasma donors.

## Author contributions

SAK, JWS, MJJ, AC, NSP conceived and designed the study. SAK, JWS, MJJ, PWJ, CCW, REC analyzed the data and performed statistical analyses. All authors reviewed, critically revised and approved the final version of the manuscript.

## Competing Interests

The authors declare no competing interests.

## Data availability

The data supporting the study findings are available within the paper.

## Funding

National Heart, Lung, and Blood Institute grant (5R35HL139854, to MJJ), the National Institute of Diabetes and Digestive and Kidney Diseases (5T32DK07352, to JWS and CCW), the Natural Sciences and Engineering Research Council of Canada (PDF-532926-2019, to SAK), the National Institutes of Health (1-F32-HL154320-01 to JWS), the National Institute of Allergy and Infectious Disease (R21 AI145356 and R21 AI152318, to DF; R01 AI1520789, to AC), and the National Heart, Lung, and Blood Institute (R01 HL059842, to AC)

## Notes

### Competing Interest Statement

The authors have declared no competing interest.

### Funding Statement

None.

### Author Declarations

All procedures accessed public information and did not require ethical review as determined by the Mayo Clinic Institutional Review Board in accordance with the Code of Federal Regulations, 45 CFR 46.102, and the Declaration of Helsinki.

### Summary of Updates

Major changes: 1: Updated studies are current as of January 16th, 2021. There are now 128 studies included in the review. 2.We performed a subgroup analysis investigating the number of days from hospital admission to transfusion on the mortality reduction associated with convalescent plasma 3.We included a number needed to treat analysis, which yields ~11 patients (range: 8-16) must be transfused to avoid one death 4.We performed an assessment of bias for all controlled studies using published criteria (Cochrane and Newcastle-Ottawa Scale)

