## Supplemental Material for "The Effect of Convalescent Plasma Therapy on COVID-19 Patient Mortality: Systematic Review and Meta-analysis"

This appendix has been provided by the authors to give additional information about their work.

**Contents**

**Supplementary Tables**

**Table S1:** Risk of bias assessment for Randomized Clinical Trials.

**Table S2:** Risk of bias assessment for matched-control and dose-response studies.

**Table S3:** Mortality Rates among COVID-19 Patients.

**Table S4:** Descriptive Data for Controlled Convalescent Plasma Studies.

**Table S5:** Descriptive Data for Dose-Response Convalescent Plasma Studies.

**Table S6:** Descriptive Data for Uncontrolled Convalescent Plasma Studies.

**Supplementary Figures**

**Figure S1:** Flow chart of the study selection.

**Figure S2:** Funnel plot of the standard error and log odds ratio for each study. The angled lines define the area including the 95% CI of the log odds ratio and the vertical line defines the middle of the funnel at the mean log odds ratio. Visual inspection of the funnel plot shows that one study fall below 95% CI and two studies are above 95% CI.

**Table S1. Cochrane Risk of Bias Assessment for Randomized Clinical Trials.**

| | Domain 1. Randomization process | | | | Domain 2. Deviations from intended interventions | | | | | | | | Domain 3. Missing outcome data | | | | |
|---|---|---|---|---|---|---|---|---|---|---|---|---|---|---|---|---|---|
| **Study** | 1.1 | 1.2 | 1.3 | 1.0 Algorithm result | 2.1 | 2.2 | 2.3 | 2.4 | 2.5 | 2.6 | 2.7 | 2.0 Algorithm result | 3.1 | 3.2 | 3.3 | 3.4 | 3.0 Algorithm result |
| Agarwal et al. | Y | Y | N | Low | Y | Y | Y | N | NA | Y | NA | Some concerns | Y | NA | NA | NA | Low |
| AlQahtani et al. | Y | Y | N | Low | Y | Y | N | NA | NA | Y | NA | Low | Y | NA | NA | NA | Low |
| Avendana-Sola et al. | Y | Y | N | Low | Y | Y | Y | N | NA | Y | NA | Some concerns | Y | NA | NA | NA | Low |
| Bajpai et al. | Y | Y | N | Low | Y | Y | N | NA | NA | Y | NA | Low | Y | NA | NA | NA | Low |
| Gharbharan et al. | Y | Y | N | Low | Y | Y | Y | N | NA | Y | NA | Some concerns | Y | NA | NA | NA | Low |
| Li et al. | Y | Y | N | Low | Y | Y | Y | N | NA | Y | NA | Some concerns | Y | NA | NA | NA | Low |
| Libster et al. | Y | Y | N | Low | N | N | NA | NA | NA | Y | NA | Low | Y | NA | NA | NA | Low |
| Rasheed et al. | Y | Y | N | Low | Y | Y | N | NA | NA | Y | NA | Low | Y | NA | NA | NA | Low |
| Ray et al. | Y | Y | N | Low | Y | Y | Y | N | NA | Y | NA | Some concerns | Y | NA | NA | NA | Low |
| Simonovich et al. | Y | Y | N | Low | N | N | NA | NA | NA | Y | NA | Low | Y | NA | NA | NA | Low |

NA, not applicable; N, no; NI, no information; Y, yes

**Table S1. Continued.**

| | Domain 4. Measurement of the outcome | | | | | | Domain 5. Selection of the reported result | | | | Domain 6. Overall Bias |
|---|---|---|---|---|---|---|---|---|---|---|---|
| **Study** | 4.1 | 4.2 | 4.3 | 4.4 | 4.5 | 4.0 Algorithm result | 5.1 | 5.2 | 5.3 | 5.0 Algorithm result | Algorithm overall Judgement |
| Agarwal et al. | N | N | Y | N | NA | Low | Y | N | N | Low | Some concerns |
| AlQahtani et al. | N | N | Y | N | NA | Low | NI | N | N | Some concerns | Some concerns |
| Avendana-Sola et al. | N | N | Y | N | NA | Low | Y | N | N | Low | Some concerns |
| Bajpai et al. | N | N | Y | N | NA | Low | Y | N | N | Low | Low |
| Gharbharan et al. | N | N | Y | N | NA | Low | Y | N | N | Low | Some concerns |
| Li et al. | N | N | Y | N | NA | Low | Y | N | N | Low | Some concerns |
| Libster et al. | N | N | N | NA | NA | Low | Y | N | N | Low | Low |
| Rasheed et al. | N | N | Y | N | NA | Low | Y | N | N | Low | Low |
| Ray et al. | N | N | Y | N | NA | Low | NI | N | N | Some concerns | Some concerns |
| Simonovich et al. | N | N | N | NA | NA | Low | Y | N | N | Low | Low |

NA, not applicable; N, no; NI, no information; Y, yes

**Table S2. Newcastle-Ottawa Risk of Bias Assessment for Controlled Studies.**

| | Domain 1. Selection | | | | |
|---|---|---|---|---|---|
| **Study** | 1.1 | 1.2 | 1.3 | 1.4 | 1.0 Total |
| Abolghasemi et al. | Selected group | Drawn from same community as exposed cohort (*) | Secure record (*) | No | 2 |
| ah Yoon et al. | Somewhat representative (*) | Drawn from same community as exposed cohort (*) | Secure record (*) | No | 3 |
| Alsharidah et al. | Somewhat representative (*) | Drawn from different source | Secure record (*) | No | 2 |
| AlShehry et al. | Selected group | Drawn from same community as exposed cohort (*) | Secure record (*) | Yes (*) | 3 |
| Altuntas et al. | Somewhat representative (*) | Drawn from same community as exposed cohort (*) | Secure record (*) | No | 3 |
| Budhiraja et al. | Selected group | Drawn from same community as exposed cohort (*) | Secure record (*) | No | 2 |
| Donato et al. | Somewhat representative (*) | Drawn from same community as exposed cohort (*) | Secure record (*) | No | 3 |
| Duan et al. | Somewhat representative (*) | Drawn from same community as exposed cohort (*) | Secure record (*) | No | 3 |
| Hegerova et al. | Somewhat representative (*) | No description of derivation of non-exposed cohort | Secure record (*) | No | 2 |
| Joyner et al. a | Somewhat representative (*) | Drawn from same community as exposed cohort (*) | Secure record (*) | Yes (*) | 4 |
| Klapholz et al. | Somewhat representative (*) | Drawn from same community as exposed cohort (*) | Secure record (*) | No | 3 |
| Klein et al. | Somewhat representative (*) | Drawn from same community as exposed cohort (*) | Secure record (*) | No | 3 |
| Liu et al. | Somewhat representative (*) | Drawn from same community as exposed cohort (*) | Secure record (*) | Yes (*) | 4 |
| Maor et al. | Somewhat representative (*) | Drawn from same community as exposed cohort (*) | Secure record (*) | Yes (*) | 4 |
| Moniuszko-Malinowska et al. | Somewhat representative (*) | Drawn from same community as exposed cohort (*) | Secure record (*) | No | 3 |
| Omrani et al. | Somewhat representative (*) | Drawn from same community as exposed cohort (*) | Secure record (*) | No | 3 |
| Perotti et al. | Somewhat representative (*) | Drawn from different source | Secure record (*) | No | 2 |
| Rogers et al. | Somewhat representative (*) | Drawn from same community as exposed cohort (*) | Secure record (*) | No | 3 |
| Salazar M. et al. | Somewhat representative (*) | Drawn from same community as exposed cohort (*) | Secure record (*) | Yes (*) | 4 |
| Salazar E. et al. | Somewhat representative (*) | Drawn from same community as exposed cohort (*) | Secure record (*) | Yes (*) | 4 |
| Xia et al. | Somewhat representative (*) | Drawn from same community as exposed cohort (*) | Secure record (*) | Yes (*) | 4 |
| Zeng et al. | Selected group | Drawn from same community as exposed cohort (*) | Secure record (*) | No | 2 |

**Table S2 Continued.**

| | Domain 2. Comparability | | |
|---|---|---|---|
| **Study** | 2.1 | 2.2 | 2.0 Total |
| Abolghasemi et al. | controls for most important factor (age) (*) | controls for disease severity (*) | 2 |
| ah Yoon et al. | controls for most important factor (age) (*) | controls for disease severity (*) | 2 |
| Alsharidah et al. | controls for most important factor (age) (*) | controls for disease severity (*) | 2 |
| AlShehry et al. | controls for most important factor (age) (*) | controls for disease severity (*) | 2 |
| Altuntas et al. | controls for most important factor (age) (*) | does not control for disease severity | 1 |
| Budhiraja et al. | controls for most important factor (age) (*) | controls for disease severity (*) | 2 |
| Donato et al. | does not control for age | controls for disease severity (*) | 1 |
| Duan et al. | controls for most important factor (age) (*) | controls for disease severity (*) | 2 |
| Hegerova et al. | controls for most important factor (age) (*) | controls for disease severity (*) | 2 |
| Joyner et al. a | controls for most important factor (age) (*) | controls for disease severity (*) | 2 |
| Klapholz et al. | controls for most important factor (age) (*) | controls for disease severity (*) | 2 |
| Klein et al. | controls for most important factor (age) (*) | controls for disease severity (*) | 2 |
| Liu et al. | controls for most important factor (age) (*) | controls for disease severity (*) | 2 |
| Maor et al. | does not control for age | does not control for disease severity | 0 |
| Moniuszko-Malinowska et al. | does not control for age | does not control for disease severity | 0 |
| Omrani et al. | controls for most important factor (age) (*) | controls for disease severity (*) | 2 |
| Perotti et al. | controls for most important factor (age) (*) | does not control for disease severity | 1 |
| Rogers et al. | controls for most important factor (age) (*) | controls for disease severity (*) | 2 |
| Salazar M. et al. | does not control for age | controls for disease severity (*) | 1 |
| Salazar E. et al. | controls for most important factor (age) (*) | controls for disease severity (*) | 2 |
| Xia et al. | controls for most important factor (age) (*) | controls for disease severity (*) | 2 |
| Zeng et al. | controls for most important factor (age) (*) | controls for disease severity (*) | 2 |

**Table S2 Continued.**

| | | | Domain 3. Outcomes | | Domain 4. Overall Bias |
|---|---|---|---|---|---|
| **Study** | 3.1 | 3.2 | 3.3 | 3.0 Total | 4.0 Total |
| Abolghasemi et al. | record linkage (*) | Yes (28+ days) (*) | complete follow-up or lost <20% to follow-up (*) | 3 | 7 |
| ah Yoon et al. | record linkage (*) | Yes (28+ days) (*) | complete follow-up or lost <20% to follow-up (*) | 3 | 8 |
| Alsharidah et al. | record linkage (*) | Yes (28+ days) (*) | complete follow-up or lost <20% to follow-up (*) | 3 | 7 |
| AlShehry et al. | record linkage (*) | Yes (28+ days) (*) | complete follow-up or lost <20% to follow-up (*) | 3 | 8 |
| Altuntas et al. | record linkage (*) | Yes (28+ days) (*) | complete follow-up or lost <20% to follow-up (*) | 3 | 7 |
| Budhiraja et al. | record linkage (*) | Yes (28+ days) (*) | complete follow-up or lost <20% to follow-up (*) | 3 | 7 |
| Donato et al. | record linkage (*) | No | complete follow-up or lost <20% to follow-up (*) | 3 | 7 |
| Duan et al. | record linkage (*) | No | complete follow-up or lost <20% to follow-up (*) | 2 | 7 |
| Hegerova et al. | record linkage (*) | No | complete follow-up or lost <20% to follow-up (*) | 2 | 6 |
| Joyner et al. a | record linkage (*) | Yes (28+ days) (*) | complete follow-up or lost <20% to follow-up (*) | 3 | 9 |
| Klapholz et al. | record linkage (*) | No | complete follow-up or lost <20% to follow-up (*) | 2 | 7 |
| Klein et al. | record linkage (*) | Yes (28+ days) (*) | complete follow-up or lost <20% to follow-up (*) | 3 | 8 |
| Liu et al. | record linkage (*) | Yes (28+ days) (*) | complete follow-up or lost <20% to follow-up (*) | 3 | 9 |
| Maor et al. | record linkage (*) | No | complete follow-up or lost <20% to follow-up (*) | 2 | 6 |
| Moniuszko-Malinowska et al. | record linkage (*) | Yes (28+ days) (*) | complete follow-up or lost <20% to follow-up (*) | 3 | 6 |
| Omrani et al. | record linkage (*) | Yes (28+ days) (*) | complete follow-up or lost <20% to follow-up (*) | 3 | 8 |
| Perotti et al. | record linkage (*) | No | complete follow-up or lost <20% to follow-up (*) | 2 | 5 |
| Rogers et al. | record linkage (*) | Yes (28+ days) (*) | complete follow-up or lost <20% to follow-up (*) | 3 | 8 |
| Salazar M. et al. | record linkage (*) | Yes (28+ days) (*) | complete follow-up or lost <20% to follow-up (*) | 3 | 8 |
| Salazar E. et al. | record linkage (*) | Yes (28+ days) (*) | complete follow-up or lost <20% to follow-up (*) | 3 | 9 |
| Xia et al. | record linkage (*) | Yes (28+ days) (*) | complete follow-up or lost <20% to follow-up (*) | 3 | 9 |
| Zeng et al. | record linkage (*) | Yes (28+ days) (*) | complete follow-up or lost <20% to follow-up (*) | 3 | 7 |

**Table S3 | Mortality Rates among COVID-19 Patients**

| Study | Location | Convalescent Plasma | | |
|---|---|---|---|---|
| | | Survivor | Non-Survivor | Mortality |
| **Case-Series or Reports** | | | | |
| Abid et al. | Wisconsin, USA | 1 | 1 | 50% |
| Ahn et al. | KOR | 2 | 0 | 0% |
| Anderson et al. | Tennessee, USA | 1 | 0 | 0% |
| Antony et al. | Texas, USA | 1 | 0 | 0% |
| Anupama et al. | New York, USA | 1 | 0 | 0% |
| Avanzato et al. | Montana, USA | 1 | 0 | 0% |
| Baang et al. | Michigan, USA | 1 | 0 | 0% |
| Balashov et al. | RUS | 1 | 0 | 0% |
| Bao et al. | CHN | 1 | 0 | 0% |
| Betrains et al. | BEL | 4 | 1 | 20% |
| Bhumbra et al. | Indiana, USA | 1 | 1 | 50% |
| Bobek et al. | HUN | 2 | 0 | 0% |
| Bradfute et al. | New Mexico, USA | 10 | 2 | 17% |
| Choudhury et al. | IND | 1 | 0 | 0% |
| Christensen et al. | Virginia, USA | 0 | 1 | 100% |
| Cinar et al. | TUR | 1 | 0 | 0% |
| Clark et al. | FRA | 1 | 0 | 0% |
| Diorio et al. | Pennsylvania, USA | 3 | 1 | 25% |
| Donzelli et al. | ITA | 1 | 0 | 0% |
| Dulipsingh et al. | Connecticut, USA | 20 | 19 | 49% |
| Easterlin et al. | California, USA | 1 | 0 | 0% |
| Einollahi et al. | IRN | 1 | 0 | 0% |
| Erkurt et al. | TUR | 20 | 6 | 23% |
| Ferrari et al. | ITA | 7 | 0 | 0% |
| Figlerowicz et al. | POL | 1 | 0 | 0% |
| Fisher et al. | ISR | 1 | 0 | 0% |
| Fung et al. | California, USA | 4 | 0 | 0% |
| Gazitua et al. | CHL | 161 | 31 | 16% |
| Gemici et al. | TUR | 15 | 25 | 63% |
| Gonzalez et al. | ARG | 201 | 71 | 26% |
| Hahn et al. | NOR | 1 | 0 | 0% |
| Hartman W. et al. (Translational medicine communications) | Wisconsin, USA | 27 | 4 | 13% |
| Hartman W. et al. (Clinical Oncology: Case Reports) | Wisconsin, USA | 1 | 0 | 0% |
| Hatzl et al. | AUT | 2 | 0 | 0% |
| Hovey et al. | Alabama, USA | 1 | 0 | 0% |
| Hu et al. | CHN | 7 | 0 | 0% |
| Huang et al. | CHN | 20 | 4 | 17% |
| Hueso et al. | FRA | 16 | 1 | 6% |
| Im et al. | KOR | 1 | 0 | 0% |
| Jaiswal et al. | UAE | 10 | 4 | 29% |
| Jamir et al. | IND | 1 | 0 | 0% |
| Jamous et al. | South Dakota, USA | 24 | 5 | 17% |
| Ji et al. | CHN | 8 | 0 | 0% |
| Jiang et al. | CHN | 1 | 0 | 0% |
| Jin C. et al. | CHN | 6 | 0 | 0% |
| Jin H. et al. | New York, USA | 3 | 0 | 0% |
| Joyner et al. | Minnesota, USA | 17408 | 2592 | 13% |
| Karatas et al. | TUR | 1 | 0 | 0% |
| Katz-Greenberg et al. | Pennsylvania, USA | 4 | 0 | 0% |
| Kong et al. | CHN | 1 | 0 | 0% |
| Lancman et al. | New York, USA | 1 | 0 | 0% |
| Lima et al. | New York, USA | 2 | 0 | 0% |
| Luetkens et al. | Maryland, USA | 1 | 0 | 0% |
| London et al. | FRA | 1 | 0 | 0% |
| Lubnow et al. | DEU | 1 | 0 | 0% |
| Malsy et al. | DEU | 1 | 0 | 0% |
| Martinez-Resendez et al. | MEX | 8 | 0 | 0% |
| Mehta et al. | New York, USA | 1 | 1 | 50% |
| Milosevic et al. | SRB | 1 | 0 | 0% |
| Mira et al. | ESP | 1 | 0 | 0% |
| Moniuszko-Malinowska et al. | POL | 1 | 0 | 0% |
| Moore et al. | Connecticut, USA | 1 | 0 | 0% |
| Naeem et al. | Rhode Island, USA | 3 | 0 | 0% |
| Niu et al. | Louisiana, USA | 1 | 0 | 0% |
| Olivares-Gazca et al. | MEX | 8 | 2 | 20% |
| Pal et al. | Louisiana, USA | 6 | 0 | 0% |
| Peng et al. | CHN | 1 | 0 | 0% |
| Ragab et al. | EGY | 1 | 0 | 0% |
| Rahman et al. | New York, USA | 10 | 3 | 23% |
| Rizvi et al. | Michigan, USA | 1 | 0 | 0% |
| Rodriguez et al. | Georgia, USA | 1 | 0 | 0% |
| Salazar E. et al. | Texas, USA | 24 | 1 | 4% |
| Schwartz et al. | North Carolina, USA | 1 | 0 | 0% |
| Shankar et al. | IND | 1 | 0 | 0% |

| | | | | |
|---|---|---|---|---|
| Shen et al. | CHN | 5 | 0 | 0% |
| Szwebel et al. | FRA | 1 | 0 | 0% |
| Tan et al. | CHN | 1 | 0 | 0% |
| Tremblay et al. | New York, USA | 14 | 10 | 42% |
| Trimarchi et al. | ARG | 1 | 0 | 0% |
| Van Damme et al. | BEL | 1 | 0 | 0% |
| van Oers et al. | Texas, USA | 1 | 0 | 0% |
| Vlachogianni et al. | GRC | 0 | 1 | 100% |
| Wang M. et al. | CHN | 2 | 3 | 60% |
| Wang B. et al. | New York, USA | 1 | 0 | 0% |
| Wei et al. | CHN | 2 | 0 | 0% |
| Wright et al. | Texas, USA | 1 | 0 | 0% |
| Xu et al. | CHN | 1 | 0 | 0% |
| Yang et al. | CHN | 0 | 1 | 100% |
| Ye et al. | CHN | 6 | 0 | 0% |
| Yi et al. | Texas, USA | 1 | 0 | 0% |
| Yokoyama et al. | BRA | 93 | 11 | 11% |
| Zeng H. et al. | CHN | 8 | 0 | 0% |
| Zhang B. et al. | CHN | 4 | 0 | 0% |
| Zhang L. et al. (Turkish Journal of Haematology) | CHN | 1 | 0 | 0% |
| Zhang L et al. (Military Medical Research) | CHN | 2 | 0 | 0% |
| Zhang L. et al. (Aging) | CHN | 1 | 0 | 0% |
| *Case Series or Reports Total* | | 18234 | 2802 | 13% |
| *Case Series or Reports Total* [a] | | 826 | 210 | 20% |

[a] excluding Joyner et al. (n = 20,000)

**Table S4 | Descriptive Data for Controlled Convalescent Plasma Studies**

| | Convalescent Plasma | | | | | | Control | | | |
|---|---|---|---|---|---|---|---|---|---|---|
| | *n* | Women (%) | Age (years) | Mechanical Ventilation (%) | Time from admission to transfusion (days) | Follow-up (days) | *n* | Women (%) | Age (years) | Mechanical Ventilation (%) |
| **Randomized Clinical Trials** | | | | | | | | | | |
| Avendano-Sola et al. | 38 | 47 | 61 | 0 | 3 | 29 | 43 | 44 | 60 | 0 |
| Rasheed et al. | 21 | 43 | 56 | 81 | 4+ | 30 | 28 | 43 | 48 | 57 |
| Gharbharan et al. | 43 | 33 | 63 | 19 | 2 | 30 | 43 | 23 | 61 | 12 |
| AlQahtani et al. | 20 | 15 | 53 | 0 | - | 28 | 20 | 25 | 51 | 0 |
| Libster et al. | 80 | 42 | 76 | 0 | 3 | 25 | 80 | 42 | 78 | 0 |
| Li et al. | 52 | 48 | 70 | 27 | 15 | 28 | 51 | 35 | 69 | 22 |
| Ray et al. | 40 | 25 | 61 | 0 | 4 | 30 | 40 | 33 | 61 | 0 |
| Simonovich et al. | 228 | 29 | 63 | 0 | 8[a] | 30 | 105 | 39 | 62 | 0 |
| Agarwal et al. | 235 | 25 | 52 | 8 | 4 | 28 | 229 | 23 | 52 | 9 |
| Bajpai et al. | 14 | 21 | 48 | 0 | 4+ | 28 | 15 | 27 | 48 | 0 |
| **Matched-Control Studies** | | | | | | | | | | |
| Duan et al. | 10 | 40 | 53 | 30 | 6 | - | 10 | 40 | 53 | - |
| Perotti et al. | 46 | 39 | 63 | 15 | 14[a] | 7 | 23 | - | - | - |
| Omrani et al. | 40 | 15 | 48 | 20 | 10[a] | 28 | 40 | 12 | 56 | 30 |
| Hegerova et al. | 20 | - | 60 | 35 | 2 | 14 | 20 | - | - | - |
| Alsharidah et al. | 135 | 15 | 54 | 4 | 1 | 30 | 223 | 22 | 54 | 2 |
| Zeng Q. et al. | 6 | 17 | 62 | 83 | 21[a] | - | 15 | 27 | 73 | 87 |
| Donato et al. | 47 | 53 | 59 | 6 | 8[a] | 30 | 1340 | - | - | - |
| Salazar M. et al. (ARG) | 868 | 31 | 56 | 21 | - | 28 | 2298 | 33 | 64 | 22 |
| Liu et al. | 39 | 36 | 55 | 10 | 7 | 14 | 156 | 29 | 54 | - |
| Salazar E. et al. (USA) | 152 | 42 | 51 | 7 | 2 | 60 | 269 | 42 | 51 | 7 |
| Xia et al. | 138 | 44 | 65 | 1 | 10 | 14 | 1430 | 50 | 63 | 0.2 |
| Abolghasemi et al. | 115 | 42 | 54 | 0 | 3 | 30 | 74 | 50 | 57 | 0 |
| AlShehry et al. | 40 | 18 | 50 | 63 | - | 30 | 124 | 16 | 53 | 64 |
| Budhiraja et al. | 333 | 20 | 60 | 28 | - | 28 | 361 | 28 | 59 | 31 |
| ah Yoon et al. | 73 | 44 | 67 | 12 | 3 | 28 | 73 | 36 | 66 | 12 |
| Rogers et al. | 64 | 42 | 61 | 11 | 7[a] | 28 | 177 | 46 | 61 | 12 |
| Altuntas et al. | 888 | 31 | 60 | 49 | 5+ | 17 | 888 | 29 | 61 | 55 |
| Klapholz et al. | 47 | 38 | 58 | 19 | 5 | 7 | 47 | 38 | 58 | 19 |
| Klein et al. | 34 | 32 | 55 | 82 | - | 30 | 34 | 32 | 57 | 82 |
| Moniuszko-Malinowska et al. | 55 | 36 | 60 | 11 | 7[a] | 30 | 715 | 47 | 52 | 4 |

[a] denotes time from symptom onset to transfusion
- denotes data not reported

**Table S5 | Descriptive Data for Dose-Response Convalescent Plasma Studies**

| | Convalescent Plasma Higher Titer | | | | | | Convalescent Plasma Lower Titer | | | |
|---|---|---|---|---|---|---|---|---|---|---|
| | *n* | Women (%) | Age (years) | Mechanical Ventilation (%) | Time from admission to transfusion (days) | Follow-up (days) | *n* | Women (%) | Age (years) | Mechanical Ventilation (%) |
| **Dose-Response Studies** | | | | | | | | | | |
| Joyner et al. | 515 | 39 | 61 | 31 | 5 | 30 | 561 | 36 | 62 | 33 |
| Maor et al. | 19 | ~29 | ~64 | ~57 | ~10 | 14 | 30 | - | - | - |

- denotes data not reported

| Table S6 \| Descriptive Data for Uncontrolled Convalescent Plasma Studies | | | | | | |
|---|---|---|---|---|---|---|
| | Convalescent Plasma | | | | | |
| | *n* | Women (%) | Age (years) | Mechanical Ventilation (%) | Time from admission to transfusion (days) | Follow-up (days) |
| **Case Series or Reports** | | | | | | |
| Abid et al. | 2 | 0 | 82 | 50 | 8a | 5 |
| Ahn et al. | 2 | 50 | 69 | 100 | - | 15 |
| Anderson et al. | 1 | 100 | 35 | 0 | - | 10 |
| Antony et al. | 1 | 0 | 63 | 100 | 14a | RH |
| Anupama et al. | 1 | 0 | 66 | 100 | 5 | 30 |
| Avanzato et al. | 1 | 100 | 71 | 0 | 70a | 35 |
| Baang et al. | 1 | 0 | 60 | 0 | 31 | 100 |
| Balashov et al. | 1 | 100 | 9 months | 0 | 47a | - |
| Bao et al. | 1 | 0 | 38 | 100 | - | 31 |
| Betrains et al. | 5 | 100 | 37 | 0 | 56a | 118 |
| Bhumbra et al. | 2 | 0 | <18 | 100 | 10 | 7 |
| Bobek et al. | 2 | 0 | 66 | 100 | - | 14 |
| Bradfute et al. | 12 | 33 | 52 | 58 | 4 | 9 |
| Choudhury et al. | 1 | 0 | 52 | 0 | 8a | 6 |
| Christensen et al. | 1 | 100 | 64 | 100 | - | - |
| Cinar et al. | 1 | 0 | 55 | 0 | - | 11 |
| Clark et al. | 1 | 100 | 76 | 0 | 50 | 36 |
| Diorio et al. | 4 | - | 14-18 | 75 | 12a | 27 |
| Donzelli et al. | 1 | 100 | 34 | 100 | 2 | 42 |
| Dulipsingh et al. | 39 | - | >18 | - | - | 7 |
| Easterlin et al. | 1 | 100 | 22 | 100 | 2 | 25 |
| Einollahi et al. | 1 | 0 | 42 | 100 | 12 | 32 |
| Erkurt et al. | 26 | 31 | 67 | 65 | 14 | 7 |
| Ferrari et al. | 7 | 14 | 60 | 14 | 8a | 30 |
| Figlerowicz et al. | 1 | 100 | 6 | 0 | 37a | 21 |
| Fisher et al. | 1 | 100 | 65 | 0 | - | - |
| Fung et al. | 4 | 25 | 53 | 25 | 6a | 29 |
| Gazitua et al. | 192 | 30 | 59 | 41 | - | 28 |
| Gemici et al. | 40 | 27 | 58 | 83 | 5a | 13 |
| Gonzalez et al. | 272 | 28 | 53 | - | - | 28 |
| Hahn et al. | 1 | 0 | ~70 | 100 | 31 | 32 |
| Hartman W. et al. (Translational medicine communications) | 31 | 32 | - | 32 | - | 7 |
| Hartman W. et al. (Clinical Oncology: Case Reports) | 1 | 0 | 35 | 0 | - | 61 |
| Hatzl et al. | 2 | 0 | 54 | 100 | 17a | 42 |

| | | | | | | |
|---|---|---|---|---|---|---|
| Heueso et al. | 17 | 71 | 58 | 12 | - | 7 |
| Hovey et al. | 1 | 0 | 26 | 0 | 11 | 3 |
| Hu et al. | 7 | 43 | 64 | 14 | 23 | 20 |
| Huang et al. | 24 | 46 | 67 | 17 | 18 | 6 |
| Im et al. | 1 | 0 | 68 | 100 | - | 23 |
| Jaiswal J. et al. | 14 | 21 | 51 | 100 | 9a | 28 |
| Jamir et al. | 1 | 0 | 49 | 0 | 2a | 21 |
| Jamous et al. | 29 | 24 | 58 | 68 | - | 28 |
| Ji et al. | 8 | 13 | 70 | 0 | 8 | 7 |
| Jiang et al. | 1 | 100 | 70 | 0 | 4 | 26 |
| Jin et al. | 6 | 33 | 61 | 50 | - | 60 |
| Jin et al. | 3 | 0 | 24 | 0 | - | 31 |
| Joyner et al. | 20,000 | 39 | 63 | 36 | 5 | 7 |
| Karatas et al. | 1 | 0 | 61 | 0 | 40a | 38 |
| Katz-Greenberg et al. | 4 | 75 | 52 | 50 | - | - |
| Kong et al. | 1 | 0 | 100 | 0 | - | 13 |
| Lancman et al. | 1 | 100 | 55 | 0 | 81a | ~5 |
| Lima et al. | 2 | 0 | 65 | 100 | - | RH |
| Leutkens et al. | 1 | 100 | 72 | 0 | 3a | 2 |
| London et al. | 1 | 100 | 41 | 0 | 71a | 4 |
| Lubnow et al. | 1 | 100 | 21 | 0 | 15 | 25 |
| Malsy et al. | 1 | 100 | 53 | 0 | ~90a | ~50 |
| Martinez-Resendez et al. | 8 | 25 | 57 | 63 | - | 23 |
| Mehta et al. | 2 | 0 | 58 | 100 | - | - |
| Milosevic et al. | 1 | 0 | 35 | 0 | 15a | ~50 |
| Mira et al. | 1 | 0 | 39 | 0 | 23a | 7 |
| Moniuszko-Malinowska et al. | 1 | 100 | 63 | 0 | 9 | 14 |
| Moore et al. | 1 | 100 | 63 | 0 | 88a | 2 |
| Naeem et al. | 3 | 67 | 36 | 67 | 3a | 13 |
| Niu et al. | 1 | 100 | 53 | 0 | 4a | 10 |
| Olivares-Gazca et al. | 10 | 0 | 52 | 50 | - | 8 |
| Pal et al. | 6 | 50 | 59 | 0 | 5a | 12 |
| Peng et al. | 1 | 100 | 66 | 100 | - | 26 |
| Ragab et al. | 1 | 0 | 72 | 0 | 6 | 33 |
| Rahman et al. | 13 | 38 | 51 | 0 | 8a | - |
| Rizvi et al. | 1 | 0 | 51 | 100 | 28 | 60 |
| Rodriguez Z. et al. | 1 | 100 | 9 weeks | 100 | 28a | 17 |
| Salazar E. et al. | 25 | 56 | 51 | 48 | 2 | 11 |
| Schwartz et al. | 1 | 100 | 5 | 100 | 6a | 21 |
| Shankar et al. | 1 | 100 | 4 | 0 | 8a | 10 |

| | | | | | | |
|---|---|---|---|---|---|---|
| Shen et al. | 5 | 40 | 65 | 100 | - | 47 |
| Szwebel et al. | 1 | 0 | - | 0 | 64 | 90 |
| Tan et al. | 1 | 0 | ~45 | 0 | 36 | 6 |
| Tremblay et al. | 24 | 42 | 69 | 13 | - | 9 |
| Trimarchi et al. | 1 | 0 | 24 | 0 | 9a | 5 |
| Van Damme et al. | 1 | 0 | 37 | 100 | 20 | 33 |
| van Oers et al. | 1 | 0 | <1 | 0 | 6 | 60 |
| Vlachogianni et al. | 1 | 0 | 66 | 100 | 33a | 4 |
| Wang M. et al. | 5 | 60 | 56 | 100 | 37a | 7 |
| Wang B. et al. | 1 | - | - | - | - | RH |
| Wei et al. | 2 | 0 | 66 | 0 | 39 | 18 |
| Wright et al. | 1 | 0 | 54 | 0 | 24a | 7 |
| Xu et al. | 1 | 0 | 65 | 100 | - | 11 |
| Yang et al. | 1 | 0 | 66 | 100 | 15 | 12 |
| Ye et al. | 6 | 50 | 58 | 0 | - | 25 |
| Yi et al. | 1 | - | - | 100 | - | RH |
| Yokoyama et al. | 104 | 29 | 64 | 35 | 2 | 14 |
| Zeng H. et al. | 8 | 50 | 65 | 13 | 15 | 11 |
| Zhang B. et al. | 4 | 50 | 57 | 100 | - | 38 |
| Zhang L. et al. (Turkish Journal of Haematology) | 1 | 100 | 46 | 0 | 5a | 9 |
| Zhang L et al. (Military Medical Research) | 2 | 100 | 60 | 0 | 20 | 8 |
| Zhang L. et al. (Aging) | 1 | 100 | 64 | 100 | 17 | 11 |

[a] denotes time from symptom onset to transfusion

-, data unavailable

RH, remains hospitalized

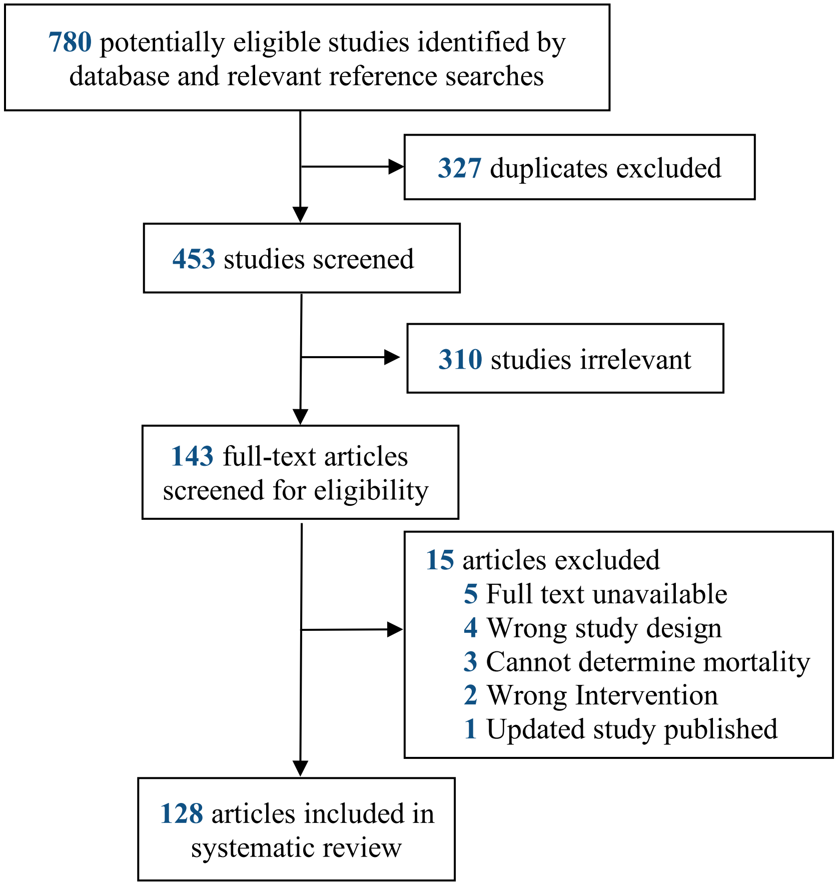


**Figure S1**. Flow chart of the study selection.

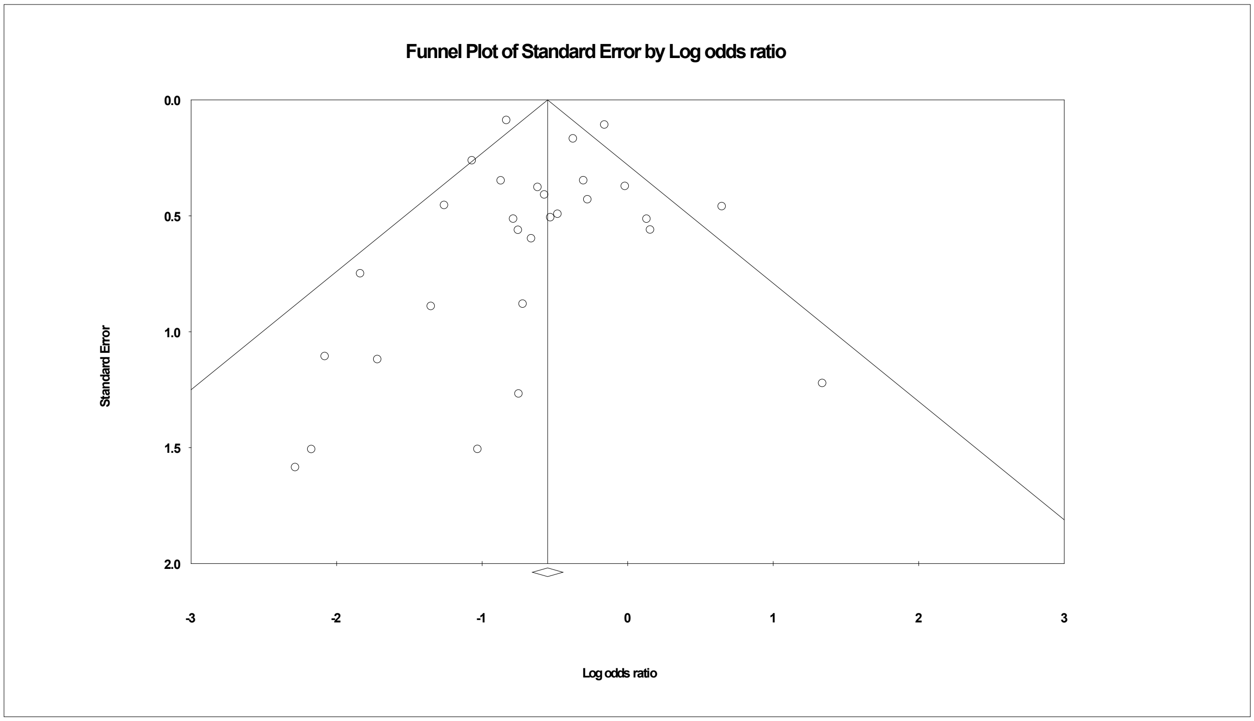


**Figure S2**. Funnel plot of the standard error and log odds ratio for each study. The angled lines define the area including the 95% CI of the log odds ratio and the vertical line defines the middle of the funnel at the mean log odds ratio. Visual inspection of the funnel plot shows that one study fall below 95% CI and two studies are above 95% CI.
